# Health Seeking Behaviour Among Men in Low-Income Settings Towards Prostate Cancer Screening Uptake, A Case Study of Mukuru Informal Setttlement, Nairobi-Kenya

**DOI:** 10.1101/2025.06.02.25328799

**Authors:** Kagwira Akule, Francis Oguya, Elizabeth Mwaniki, James Nyariki

## Abstract

**Background:** Prostate cancer screening is not a routine practice in Kenya despite of high morbidity and mortality associated with the disease. Most men seek medical attention when the disease is in an advanced stage resulting in a poor prognosis.

**Objective:** This study aimed to determine the health seeking behavior and the uptake of prostate cancer screening among men aged 30-64 years old in Mukuru informal settlement, Nairobi-Kenya.

**Methods:** The study deployed a descriptive cross-sectional utilizing both primary and secondary data. Primary data was collected through semi-structured questionnaires, key informant guides, and observation methods. Secondary data was obtained from existing facility health records, files and Ministry of Health (MoH) reporting tools for non-communicable diseases.

**Participants:** The sample size comprised 200 male respondents aged between 30-64 years old from Mukuru informal settlement in Nairobi.

**Results:** The study found that majority (80.5%) of respondents were married, had acquired formal form of education and most (50.5%) of them were self-employed while 30% were casual laborers. More than half (52.5%) resided close to a public healthcare facility and despite high (70%) awareness of prostate cancer (PCa), only a few (3.5%) reported having ever been screened for PCa. The respondents who reported being aware of the PCa screening method, whether it is manageable and known risk factors were found to be likely (p = 0.001, p = 0.002 and p = 0.001) respectively to have ever been screened for PCa. Additionally, qualitative data was collected to gain an in-depth understanding for low uptake of PCa screening.

**Conclusion:** Socio-demographic variable on education was associated with the uptake of PCa screening. The healthcare services and health-seeking behavior showed significant associations to both outcomes, “ever been screened” and willingness to be screened." Moreover, the findings from this study demonstrated that the unavailability of free PCa screening services in public healthcare facilities, insufficient information on PCa screening, Lack of knowledge on predisposing risk factors and poor health-seeking behavior among men in Mukuru informal settlement resulted in a deficient uptake of PCa screening.

**Recommendation:** Therefore, this study recommends for the local county government to partner with MoH in creating critical awareness on the importance of routine PCa screening practices by providing information resources and free screening services in order to have a well-informed population on screening benefits, harms and risks.

## INTRODUCTION

Prostate cancer (PCa) affects the male prostate gland and is increasingly becoming one of the most public health concerns and the most common malignancy seen in men worldwide [1]. The prostate gland is a male reproductive organ located just below the urinary bladder surrounding the urethra and in front of the rectum. It is a walnut-sized gland that produces seminal fluids that aid in the transportation and nourishment of sperm. Prostate cancer screening is the process of detecting a disease in the asymptomatic stage. Globally, PCa is the second most commonly identified male disease and the leading cause of male fatalities worldwide, precisely in underdeveloped countries and the Caribbean [2]. In the region, prostate cancer screening is still a contentious threat because of the diagnosis and side effects of biopsy and treatment in industrialized nations. However, according to [3], recommended men of African ethnicity who were at higher risk would benefit the most from early PCa screening. Furthermore, the importance of including informed shared decision-making during screening has been extensively accepted globally. In a collaborative decision-making process, the UN Preventive Task Force report suggests early screening of males for PCa among those who are well thought-out to be at risk especially men with a family history of the disease and of African ethnicity [4].

Kenya continues to have extremely poor screening uptake, much like other underdeveloped nations. The level of participation is still low nationally even though Kenya’s current guidelines advocate screening of men aged 40 to 69 years through an informed shared decision-making process [5]. The disease is accelerated by genetic variations based on geographic distribution. Other significant risk factors for the growth of prostatic carcinoma include androgens, aging, insufficient food, cigarette use, excessive alcohol use, obesity and ethnicity [7].

A prospective cohort study (2020-2021) that was done in the oncology department at Kenyatta National Hospital (KNH) in Nairobi-Kenya reported a mortality rate due to PCa was 4.9% annually [8]. This is due to the late diagnosis and presentation of PCa patients resulting in a significant cancer death rate in the country. In addition, insufficient Cancer Centers in the country have also attributed to poor prognosis, which is a reflection of limited access to early detection and treatment. One of the socio-demographic aspects that affects the uptake of men’s cancer screening globally is age. About 1 in 8 men during their lifetime will be diagnosed with PCa based on age, race and other factors. For instance, about 6 in 10 prostate cancer patients are first diagnosed at the age of 65 or older, and the average age is 67 years old [9]. According to [10] report recommends PCa screening according to risk factors as follows; men aged 50 who are at average risk of developing PCa and have more than 10 years of life expectancy, men aged 45 with a family history of PCa and of African ancestry (high risk) and at age 40 of men with the first degree of relative diagnosed with PCa at an early age (higher risk). These results show PCa increases in men beyond the age of 40 years [10]. Although age, ethnicity, and heritage are the primary risk factors for the disease, there is evidence that men of high-class social status have an increased prostate cancer screening uptake and that their tumors are more likely to be identified early, leading to reduced fatality rates compared to low-income social status [11].

Based on the report from the [12], 50% of the world’s population is unable to access primary healthcare services. However, the Global Agenda for Universal Health Coverage conference reported that 11 million Africans live in poverty as a result of spending their household income to pay for basic medical care [13]. Based on the report from [13] primary healthcare services are not equally accessible, affordable, and available in countries in Southern Asia and Sub-Saharan Africa. The burden of prostate cancer in developing nations is mostly a result of limited access to services such as screening, prevention, and control [13].

They also noted the gap between industrialized and unindustrialized nations on PCa prevalence and mortality is primarily a result of effectual prevention and control services for PCa in these industrialized nations. The most proficient way to attain the best health outcome in low-income populations is through the provision of affordable, accessible, available and acceptable healthcare services. The level of socio-economic, socio-demographic and social barriers determines how people utilize healthcare services [14]. However, like any other underdeveloped countries, the main obstacles to receiving healthcare services depend on how accessible, available and affordable these services particularly for low-income populations.

In Kenya, the Ministry of Health launched national cancer screening guidelines (2018) that endorse shared decision-making during PCa screening among men aged 40 to 69 years. Although the rate of PCa screening remained low with majority of males seeking treatment when the disease is advanced stage has contributed to the rise in mortality due to poor prognosis and suboptimal health-seeking behavior [15]. Currently, Kenya-cancer-policy has revealed that there are insufficient cancer treatment facilities within the country and most cancer diagnostic services are only accessible at the County referral (level 5 tertiary) and National referral hospitals, which are predominantly found in urban areas [16]. Kenya’s inadequate infrastructure for cancer treatment and some cancer management options being difficult to access have contributed to the long waiting period being the reason for previously curable tumors to advance to incurable stages. Additionally, many individuals seek treatment in lower-level health facilities (levels 2, 3, and 4) where screening and diagnosis of PCa are hampered by a lack of resources and qualified staff according to the report [17].

## METHODS

### Study design

The study adopted descriptive cross-sectional study design that used a combination of qualitative and quantitative methods to describe statistical significance of the study variables and establish a relationship between these variables and as well providing in-depth information for deeper understanding and interpretation.

### Study Population

Men living in the study area between the ages of 30-64 make up the study population for this study. According to [18] census the population size of adults’ men residing in Kwa Njenga, Kwa Reuben and Viwandani was a total of 102,312 where 200 men were obtained as the sample size for this study. Men aged 30 and older were the target population since it was anticipated that most men had at least a basic understanding of health issues and were therefore aware of prostate cancer.

### Study Area

The study was conducted in the second largest informal settlement known as Mukuru after Kibera in Nairobi County in Kenya. Mukuru informal settlement is situated near Nairobi’s industrial zone approximately 7km southeast of the central business zone and It stretches along Nairobi industrial area of geographical coordinate of latitude 1°19’45"S and longitude 36°52’21"E. Mukuru informal settlement consists of the following settlements; Mukuru kwa Njenga Mukuru kwa Reuben, Viwandani, Mukuru Kayaba, Fauta Nyayo and Mariguini with an estimated population exceeding 700,000. Mukuru informal settlement it’s in Embakasi south and Makadara constituencies [18]. Mukuru residents’ livelihood is characterized by informal economic and labor force contributing to informal jobs with low wages and job insecurity [18].

### Study variables

The study variables employed for this research are illustrated in the conceptual framework in Figure 1.1. This includes; independent (predictor) variables which are socio-demographic factors, healthcare services and health-seeking behavior in influencing the uptake of PCa screening. On the PCa screening uptake, “Ever been screened” and “Willingness to be screened” in the future were two of the dependent (outcome) variables in this study. Utilization of medical services was based on availability, accessibility, affordability and social acceptability are intermediary (moderating) variables.

**Figure 1.1.**
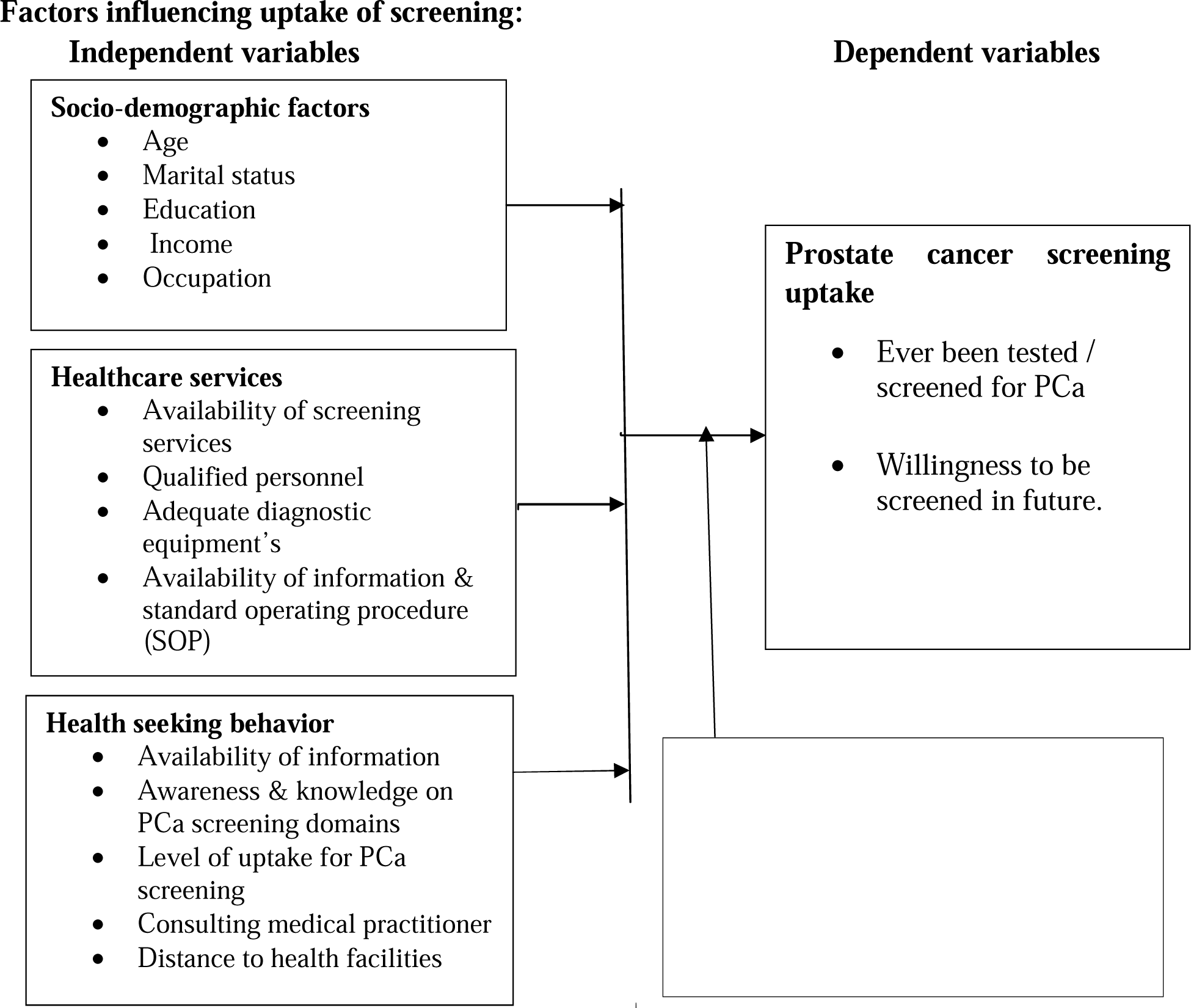
Conceptual framework

### Conceptual framework

#### Sampling Technique

Mukuru informal settlement was purposively selected because it’s considered to be a low-income setting [18], and also due to its proximity to different manufacturing industries making it a high risk area due to possible exposure to environmental pollutants and emissions from different industries. Secondly, convenient sampling was applied to select the three Mukuru Settlements (Kwa Njenga, Kwa Reuben and Viwandani) among the six Mukuru Settlements (MSs). The three Mukuru Settlements (3MSs) were grouped into clusters which was done according to administrative boundaries. A list of community units was generated from the three clusters at the chief’s camp for each study area. These units were randomly sampled from each cluster to obtain a subset sample of community units where a list of households was developed. The researcher developed a sampling frame and applied systematic random sampling to select households for each sampled community units where 200 participants were obtained based on the households heads. This was done according to population size for each cluster areas (Kwa Njenga, Kwa Rueben and Viwandani).

### Sample Size

The sample size was estimated by use of dichotomous outcome, one sample distribution [19] as follows: 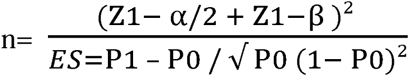 Where: n= Desired sample size

Z_1-α/2_ = level of significance at 95% (CI=1.96), α = 0.05

Z_1-β_ = 0.84 power of 80% p=0.2

ES= Effect Size

P_1_ = Estimated sample proportion of the study population which is 8.2% 0.082

P_0_ = Sample proportion of PCa screening in Nairobi County which is 4.2% 0.042 (KDHS, 2014)

q= (1- P_0_)

Therefore: ES= 0.082- 0.042 / √ 0.042(1- 0.042) = 0.199413

n= [1.96 + 0.84 / 0.199413] ^2^ = 197.16 200 (Male respondents aged 30-64 years).

### Internal Validity and Reliability

The tools are valid if they capture the data that they are intended to capture. Prior to the actual study, the instruments underwent pre-testing to ensure that was accurate and easy for the participants to understand. During the study, appropriate translation in both English and Kiswahili languages was done to remove any ambiguity and allow free flow of information between the researcher or assistants and the respondents. In order to obtain high accuracy 5% of the population frame that is eligible but not included in the final study was conducted to increase the dependability of the research apparatus. Therefore, 10 respondents aged 30-64 years old participated in the pre-test study constituting 5% of the 200 participants for this study.

### Data Collection Tool and Procedure

The data was collected using a key informant guide and observation checklist. Key informant guide was used to elicit information on the availability of screening test kits and equipment, availability of PCa screening information or guide, measures taken by the health facility in-charges to improve PCa screening policy, implementation arrangements, resources allocation and the challenges faced in provision of PCa screening services. The observation checklist was used to complement information on the availability of screening tests and equipment such as ultra-sound machines, standard operating procedure (SOP) guidelines or screening guides for PCa at their disposal and PSA screening tests.

In health-seeking behavior, the variables that were assessed include consulting a medical practitioner, information on PCa screening domains, distance to the nearest healthcare facility, payment means for healthcare services (out of pocket, insurance, family or friends support), rating of general health care service, availability of PCa screening services and level of the PCa screening uptake. In-depth interview schedules were rescheduled for both respondents and key informant interviews after obtaining voluntary informed consent. Semi-structured questionnaires, open-ended key informant guides, observational checklists and audio recordings were all used as well as documentation of field notes during the entire data collection to supplement the qualitative data.

### Data Analysis

#### Quantitative data Analysis

The data was obtained from a survey study and underwent data cleaned up, edited, coded and entered into an Excel sheet and then exported to quantitative software (SPSS version 25). The Statistical Package for Social Sciences (SPSS version 25) was used to generate descriptive and inferential findings. The results were presented in the form of tables, diagrams, frequencies and percentages for descriptive statistics. For inferential statistics to ascertain whether there was a correlation between the variables, Pearson’s Chi-square test was performed. The statistical significance was set at a p<0.05. For further analyses, binary logistic regression and Odds ratios were used to predict the probabilities of men’s intention to undergo PCa screening. The relationship between various predictor factors and the response variables "ever been screened" and "willingness to be screened" for prostate cancer was examined using binary logistic regression modelling at a significance level of less than or equal to 0.05. The odds ratio (OR) and 95% confidence interval (CI) for the variables that were determined to be significant were obtained. In this study, exposure to the predictor will increase the likelihood of the result if the OR at the 95% confidence level is greater than 1.0 conversely, exposure to the predictor would decrease the likelihood of the outcome if the OR is less than 1.0.

#### Qualitative data Analysis

The study employed deductive thematic analysis, a technique for analyzing qualitative data that entails going over a data set and checking for patterns and relationships to identify themes in response to research questions. De-identification of respondents using codes and familiarization with the data results was done. The transcript underwent an audit trail where the data was cleaned, edited and counter-checked for missed information from the audio recordings entered into an Excel sheet and then exported to the qualitative software (Nvivo Pro 12). The open-ended data was coded and categorized into words (nodes) and short phrases then thematic analysis was used to transcribe into emerging themes, ideas, patterns and relationships. The results were summarized in response to research questions in order to gain qualitative findings. Finally, the information was categorized and strongly expressed themes were examined using the qualitative Programme (Nvivo Pro 12).

#### Ethical Approvals

Ethical approval was sought from KNH-UoN Ethical Research Committee (KNH-EBC/RR/885) and statutory national permit to conduct the research was obtained from National Commission for Science, Technology and Innovation (NACOSTI / P / 93974 /46719). Authorization letters were obtained from Nairobi County Commissioner, Nairobi Metropolitan Health Services and Ministry of Education. Participant’s autonomy and privacy were maintained throughout the study and written consent was sought following the explanation of the purposes, benefits, risks and voluntary participation.

## RESULTS

### Socio-demographic characteristics of the respondents and prostate cancer screening

According to the findings, most respondents (28.5%) were between the age group of 30-34 years old, followed by those between the ages of 35 to 44 (18% and 17.5%) respectively. Those between the age group of 60-64 years old (6.5%) and 50-54 years old (6%) made up the minority. The majority of respondents were married (80.5%), only a few reported that they were single (9%) and the rest were either separated, divorced, or widower. The majority (99%) of the respondents had acquired formal education where 46% reported that the highest level of education attained was primary education and only a few (1%) reported having never schooled.

Monthly household income was collected from participants and most (25% and 27%) reported earning monthly income below KES.10,000 and KES.15,000 respectively with only a few (1%) of respondents reporting earnings above KES.30,000 per month. It was found that the majority (50.5%) of respondents were self-employed while 30% were casual laborers and only a few reported that they were supported by their family or spouses (3% and 1%) respectively. The uptake of PCa screening was very low in all the categories but with the majority (88%) willing to be screened in the future only a few (8.5%) of the respondents were not willing to uptake PCa screening as shown in Table 1

**Table 1:**
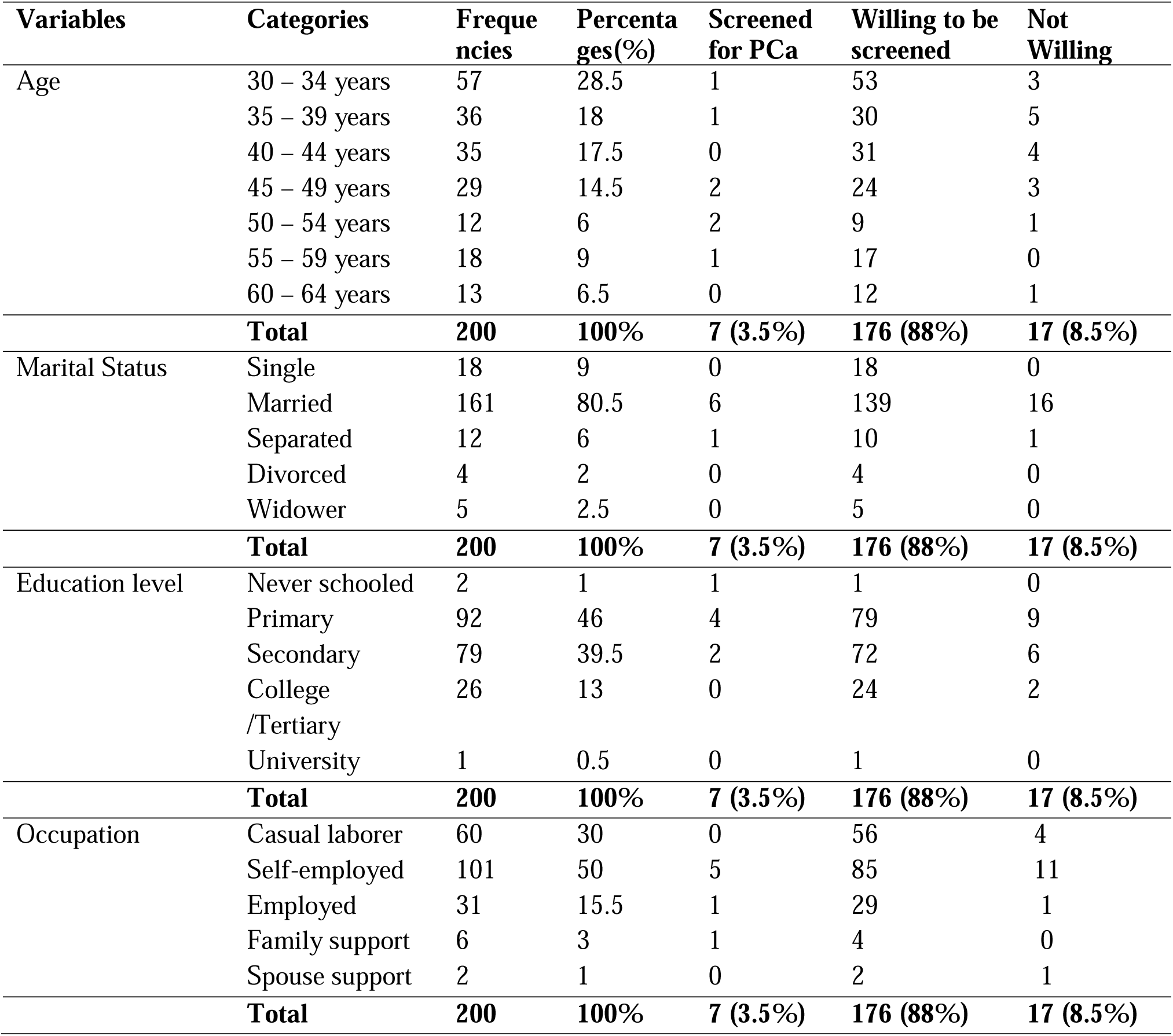
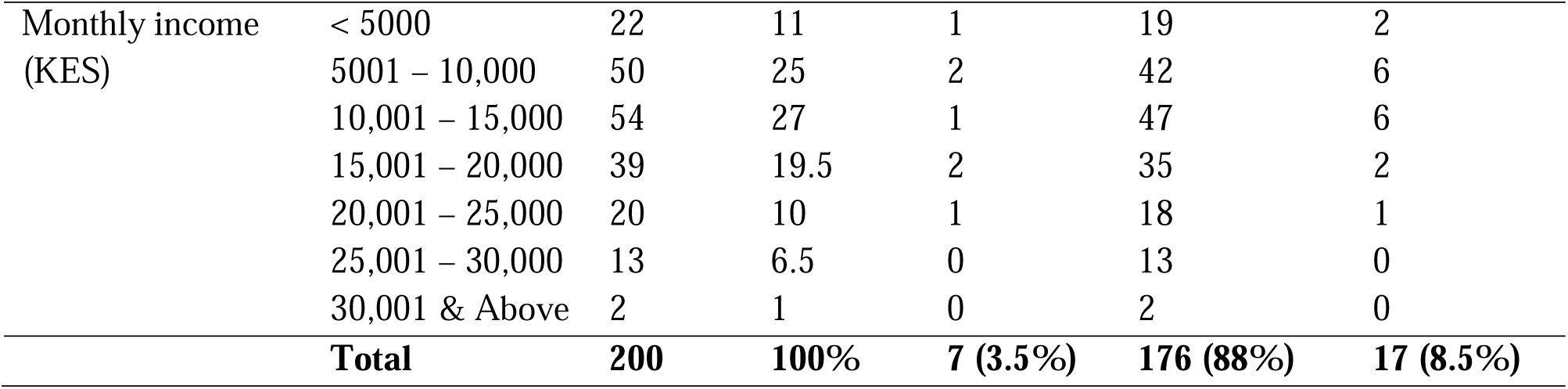
Socio-demographic characteristic and PCa screening uptake among men aged 30-64 years old from Mukuru informal settlement in Kenya.

### Association between Socio-demographic factors and the uptake of PCa screening

Level of education was observed to be associated with screening status for prostate cancer at (p=0.007) among men in Mukuru informal settlements. The acceptance of PCa screening that is being tested or desire to be tested in the future was not influenced by other factors such as age, marital status, level of income, or type of occupation (p=0.139, p=0.766, p=0.202 & p=0.958) respectively. As indicated in Table 2 below, there was no correlation between willingness to be screened with age, marital status, education, occupation, or income since the majority (96.5%) had never gone for screening by the time the study was conducted while 64.5% of respondents were not aware of PCa screening.

**Table 2.**
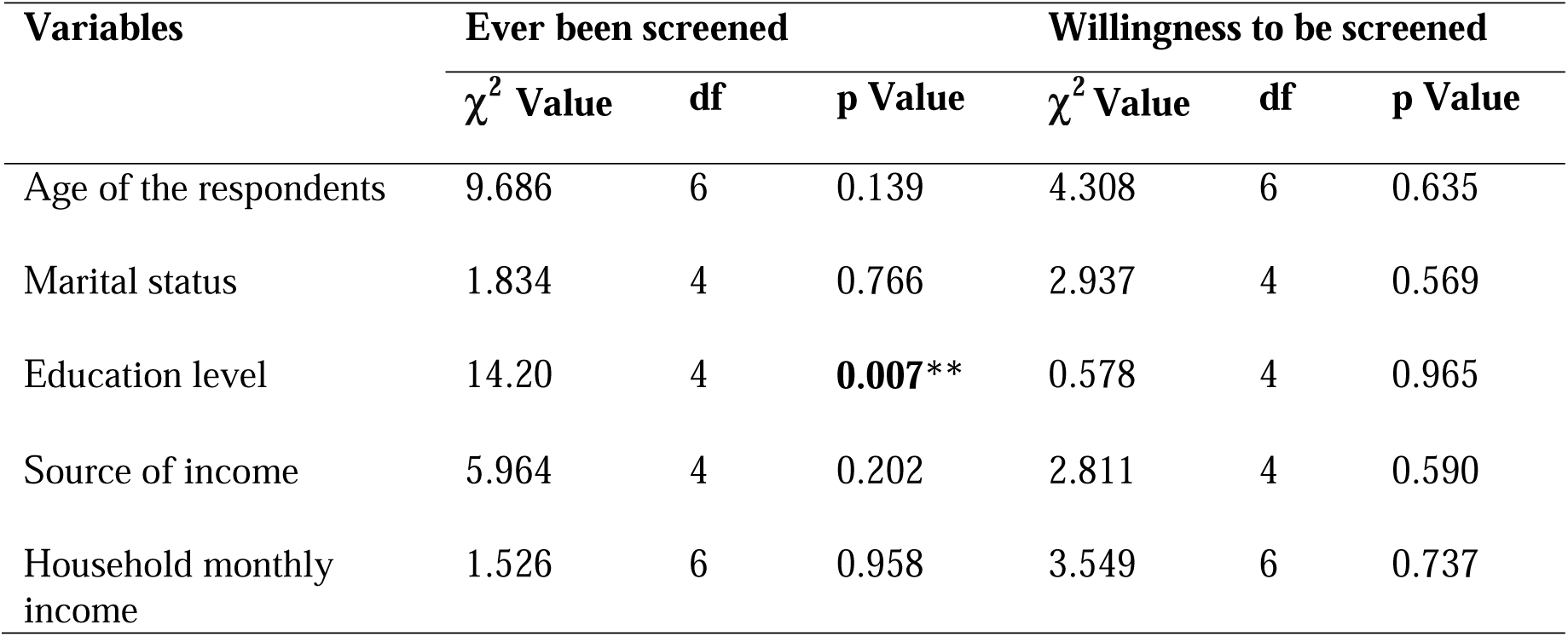
Association of socio-demographic factors with PCa screening uptake among men aged 30-64 years old from Mukuru informal settlement in Kenya.

| Variables | Ever been screened |  |  | Willingness to be screened |  |  |
| --- | --- | --- | --- | --- | --- | --- |
| | $\chi^2$ Value | df | p Value | $\chi^2$ Value | df | p Value |
| Age of the respondents | 9.686 | 6 | 0.139 | 4.308 | 6 | 0.635 |
| Marital status | 1.834 | 4 | 0.766 | 2.937 | 4 | 0.569 |
| Education level | 14.20 | 4 | **0.007\*\*** | 0.578 | 4 | 0.965 |
| Source of income | 5.964 | 4 | 0.202 | 2.811 | 4 | 0.590 |
| Household monthly income | 1.526 | 6 | 0.958 | 3.549 | 6 | 0.737 |

**Table 3.** Association of healthcare services and health seeking behavior on PCa screening uptake among men aged 30-65 years old in Mukuru informal settlement.

| Variables | Ever been screened |  |  | Willingness to be screened |  |  |
| --- | --- | --- | --- | --- | --- | --- |
| | $\chi^2$ Value | df | p Value | $\chi^2$ Value | df | p Value |
| Distance to the nearest healthcare facility | 2.699 | 2 | 0.259 | 2.956 | 2 | 0.228 |
| Consulting general medical practitioner | 0.463 | 2 | 0.793 | 9.666 | 2 | **0.008 \*\*** |
| Payment mode for health care services | 12.98 | 3 | **0.005 \*\*** | 12.71 | 3 | **0.005 \*\*** |
| Rating general health care services | 1.130 | 3 | 0.770 | 0.614 | 3 | 0.887 |
| Availability of PCa screening services | 0.582 | 3 | 0.901 | 1.480 | 3 | 0.687 |
| Aware of prostate cancer | 1.484 | 1 | 0.223 | 0.003 | 1 | 0.985 |
| Aware of PCa screening services | 3.109 | 1 | 0.078 | 2.575 | 1 | 0.109 |
| Aware of any PCa screening method | 83.42 | 1 | **0.001 \*\*** | 1.616 | 1 | 0.204 |
| Aware if PCa is manageable | 6.969 | 1 | **0.008 \*\*** | 8.270 | 1 | **0.004 \*\*** |
| Knowledgeable of any PCa risks factors | 19.61 | 1 | **0.001 \*\*** | 6.872 | 1 | **0.009 \*\*** |
| History of PCa in the family | 0.549 | 1 | 0.459 | 4.078 | 1 | **0.043 \*** |

(Asterisks indicate the level of significant differences: \* $p \leq 0.05$ ; \*\* $p \leq 0.001$ ; \*\*\* $p \leq 0.0001$ )

### Access to healthcare facilities in Mukuru slums

More than half (52.5%) of the respondents resided close to a public healthcare facility between a distance of 1 to 2 kilometers(Km) whereas 49.5% and 39.5% of the respondents reported a distance between 1 to 2 kilometers and less than 1 kilometer to a private healthcare facility respectively. Only a few (14.5%) reported less than 1 kilometer (Km) to a public healthcare facility and others (33%) reported more than 3 kilometers to a public facility as shown in Figure 2.

**Figure 2.**
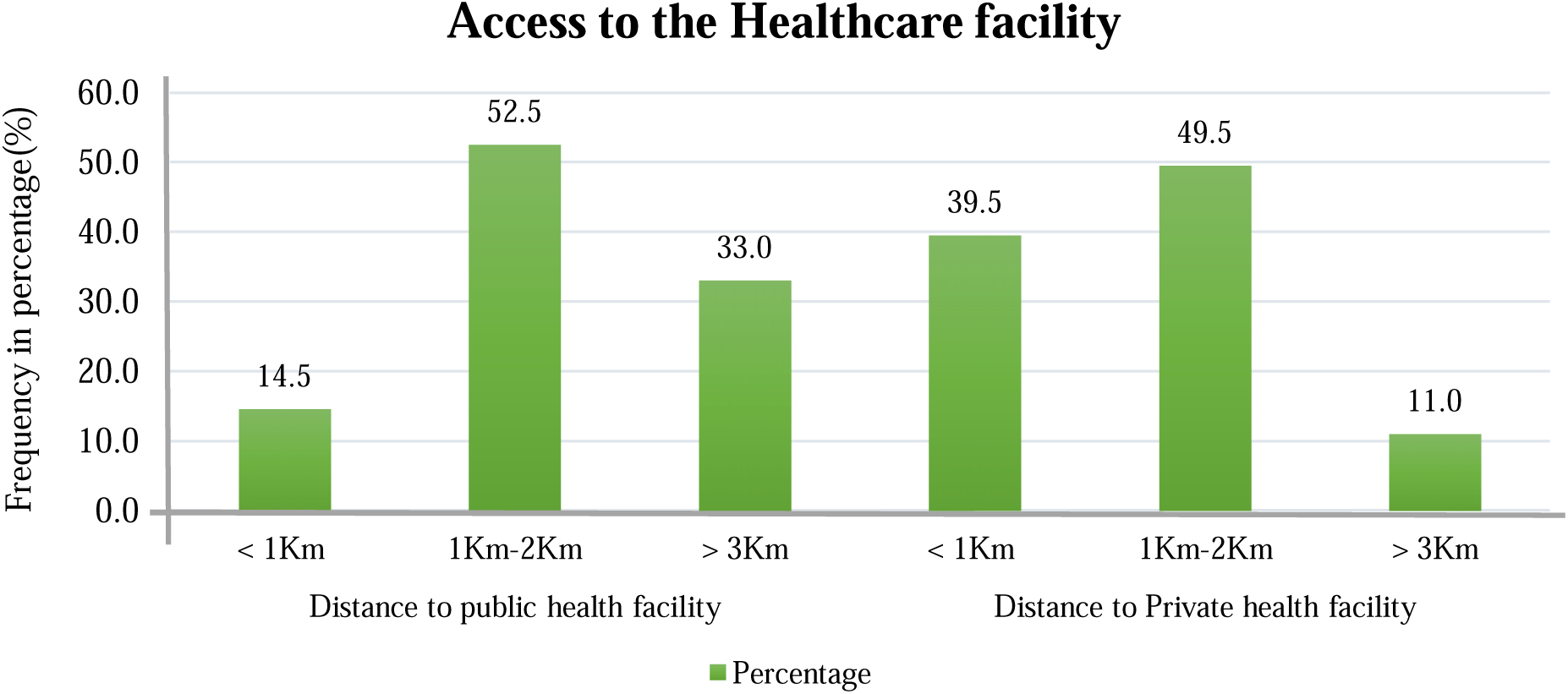
Distances to the public & private healthcare facilities in Mukuru informal settlement

**Figure 3.**
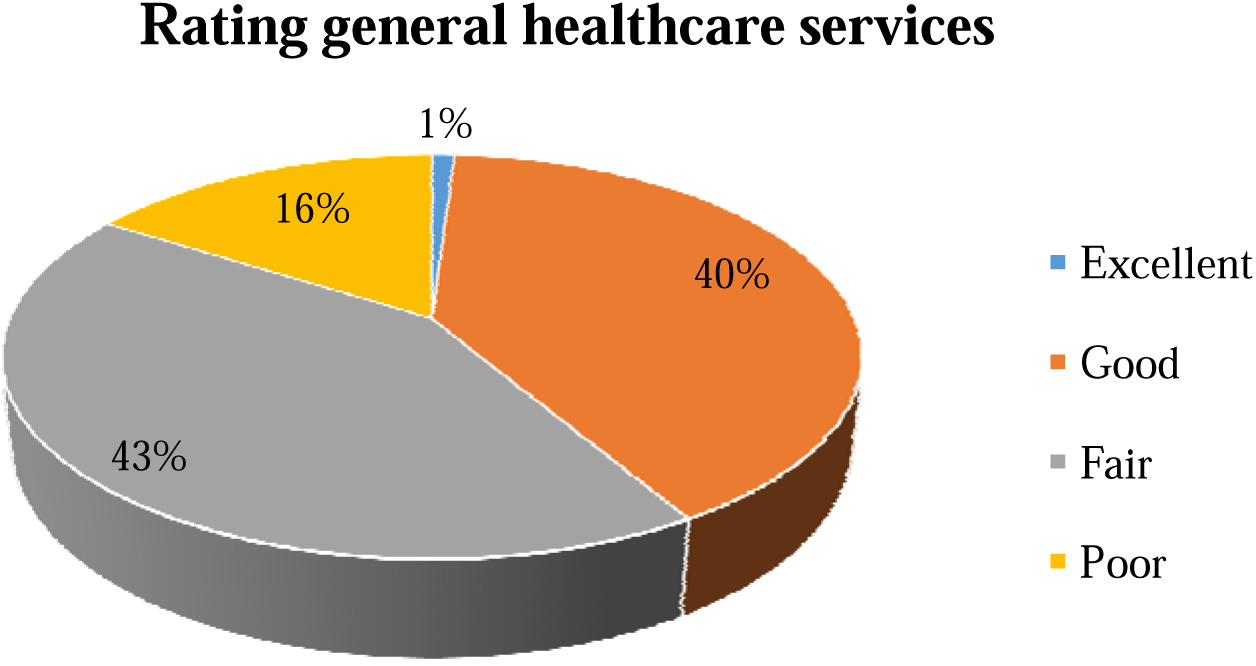
Rating of general health care services within public facilities in Mukuru

**Figure 4.**
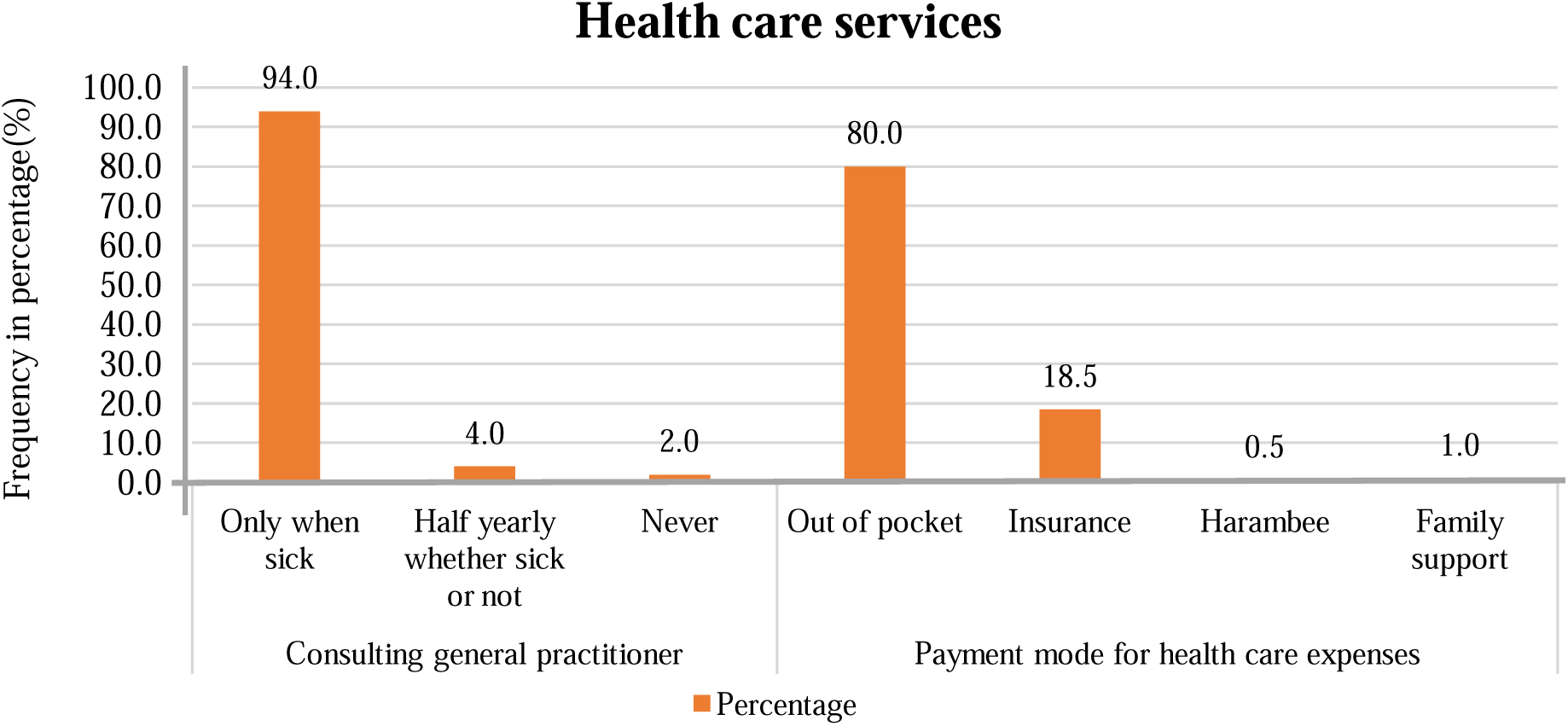
Medical consultation and payment means for health care services expenses

**Figure 5.**
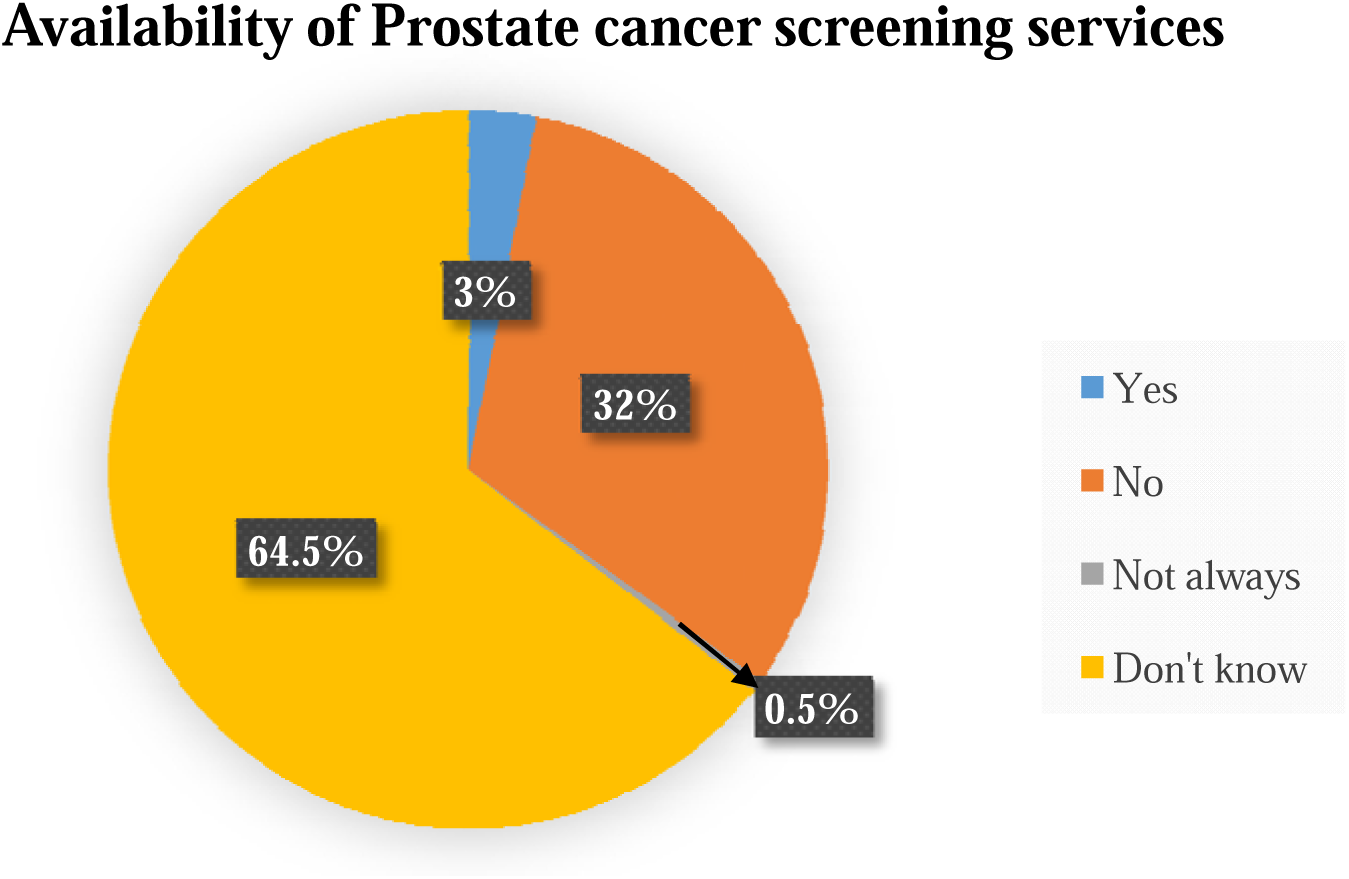
Availability of prostate cancer screening services within facilities in Mukuru

**Figure 6.**
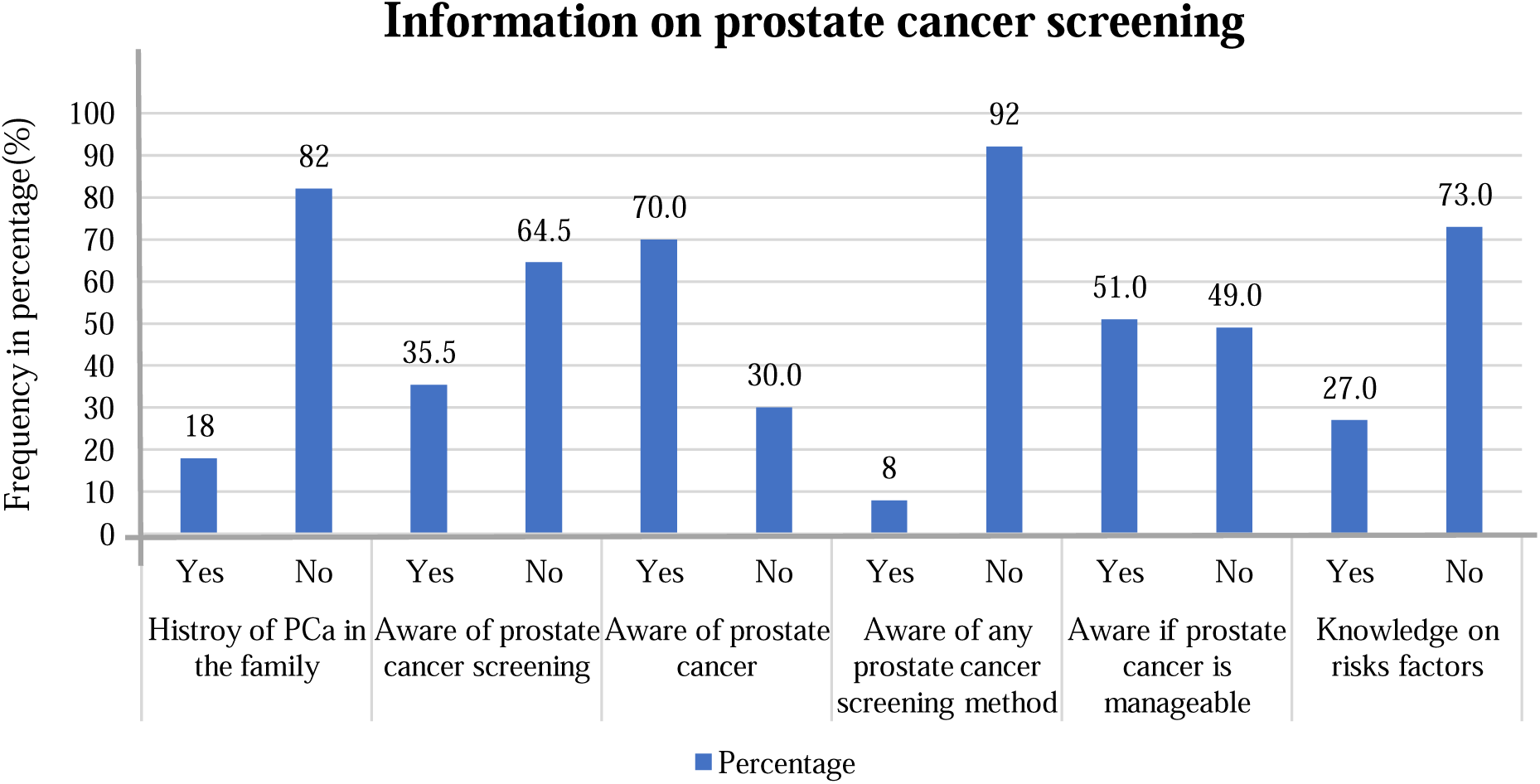
Awareness and knowledge on prostate cancer screening

**Figure 7.**
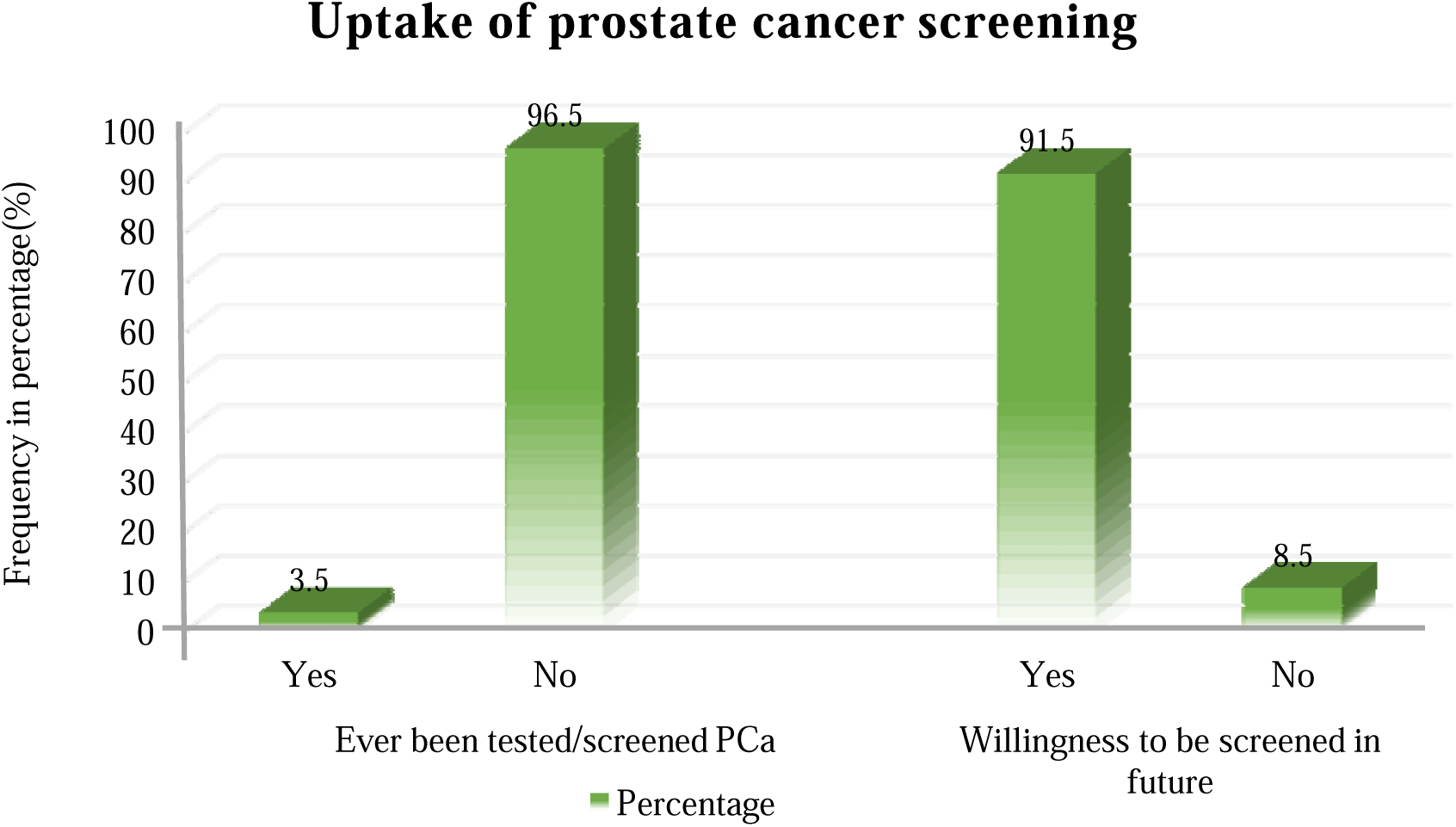
Uptake of prostate cancer screening and willingness to be screened

### Rating of general health care services in public facilities in Mukuru

Respondents rated public healthcare services as either excellent, Good, Fair or poor. Less than half (43% and 40%) of the respondents felt the services were Fair or Good respectively. Others (16%) reported that the services were poor and only a few (1%) reported excellent health care services in public healthcare facilities within Mukuru informal settlement.

### Rating general healthcare services

#### Payment means for health care expenses and medical consultation

The respondents were asked to indicate when they consulted a medical practitioner. Most of the respondents (94%) only consulted a medical practitioner when they were sick, others (4%) consulted half yearly whether sick or not and only, a few (2%) had never been sick to seek medical attention in a health facility. The respondents were asked to indicate how they paid for their healthcare expenses. The majority (80%) of the respondent indicated that they paid from their own pocket. Some (18.5%) of the respondents used insurance cover. Only a few (1%) respondents sought help from the family.

### Availability of prostate cancer screening services in healthcare facilities within Mukuru informal settlement

Only a few (3%) of the respondents reported that screening services for prostate-specific antigen (PSA) and digital rectal examination (DRE) were only available in private healthcare facilities in Mukuru informal settlements. This figure corresponds to the few (3.5%) respondents who reported had ever been screened for Prostate cancer. Additionally, 32% of the respondents reported that Prostate cancer screening was not available in public health facilities while the majority (64.5%) of the respondents were unaware of the availability of PCa screening and testing services.

### Health seeking behavior among men in Mukuru informal settlement

There was a high level of awareness about prostate cancer with 70% of the respondents reported to have heard about the disease. However, the majority (64.5% and 92%) were not aware of PCa screening neither any screening method used respectively, whereas half (51%) of them were aware that prostate cancer is manageable when detected early enough. Only a few (21%) knew predisposing risk factors of PCa and 18% reported having cases of PCa in the family.

### Level of uptake for prostate cancer screening among men in Mukuru settlements

Despite the high level of PCa awareness above in (Figure: 4.5), only (3.5%) of the respondents reported having undergone a PCa screening by the time the study was conducted. However, after learning more the majority of respondents (91.5%) who took part in the study agreed to be screened in the future at the end of the interview if these services are freely offered to their nearest healthcare facilities.

### Healthcare services and health seeking behavior influencing the uptake of PCa screening

In this study, it was found that distance to the facility, rating of general health care service and availability of PCa screening services were not associated with ever been screened or willingness to be screened. The results indicate that payment mode for health care service was significantly associated with ever been screened and willingness to be screened both at a p=0.005. On the health-seeking behavior, the variables that were assessed found that individuals who were aware of PCa screening procedures, risk factors and aware that PCa is manageable were more likely to have undergone screening (p=0.001, p=0.001 and p=0.008) respectively. There was no significant association between consulting a general medical practitioner, awareness of PCa, awareness of PCa screening and family history of PCa with ever been screened for PCa (p=0.793, p=0.223, p=0.078 and p=0.459) respectively.

Willingness to be screened for PCa among men in Mukuru was significantly associated with how frequently they consulted a medical practitioner (p=0.008). However, being aware of the disease, being aware of PCa screening and awareness of PCa screening methods were not associated with the uptake (p=0.985, p=0.104 and p=0.204) respectively. Awareness that PCa is manageable, knowledge of risk factors and reported cases of cancer in the family were significantly associated with willingness to be screened in the future (p=0.004, p=0.009 and p=0.043) respectively.

### Association between healthcare and health seeking behavior with prostate cancer screening

Binary logistic regression was carried out to assess the association between healthcare and health-seeking behavior and uptake of prostate cancer screening. Variables that were assessed under healthcare services the study found that there was no association between healthcare services with both “ever been screened” and “willingness to be screened” for PCa in the future. However, awareness of PCa screening, awareness of any PCa screening method and awareness if PCa is manageable showed significant (p=0.024, p=0.001 and p=0.002 respectively) association between health-seeking behavior and ever been screened. Additionally, respondents who had knowledge of any PCa risk factors were significantly associated with both “ever been screened” (p=0.001) and “willingness to be screened” (p=0.020). Consulting a general medical practitioner showed statistical significance (p=0.046) in willingness to be screened for prostate cancer among men in Mukuru informal settlement. Similarly, the respondents who reported a history of PCa in the family were significantly (p=0.008) associated with willingness to be screened for prostate cancer in the future as illustrated in Table 4.

**Table 4.** Binary logistic regression analysis on healthcare and health seeking behavior among men aged 30-64 years old from Mukuru informal settlement in Kenya.

| Variables | Ever been screened |  |  | Willingness to be screened |  |  |
| --- | --- | --- | --- | --- | --- | --- |
|  | Likelihood ratio | df | p Value | Likelihood ratio | df | p Value |
| Distance to the nearest healthcare facility | 0.333 | 2 | 0.847 | 2.917 | 2 | 0.233 |
| Consulting general medical practitioner | 0.882 | 2 | 0.643 | 6.159 | 2 | **0.046 \*** |
| Payment mode for health care services | 4.219 | 3 | 0.239 | 7.570 | 3 | 0.056 |
| Rating general health care services | 1.110 | 3 | 0.775 | 0.814 | 3 | 0.846 |
| Availability of PCa screening services | 0.804 | 3 | 0.848 | 2.090 | 3 | 0.554 |
| Aware of prostate cancer | 1.408 | 1 | 0.235 | 0.000 | 1 | 0.985 |
| Aware of PCa screening services | 5.102 | 1 | **0.024 \*** | 2.397 | 1 | 0.122 |
| Aware of any PCa screening method | 38.756 | 1 | **0.001 \*\*** | 2.969 | 1 | 0.085 |
| Aware if PCa is manageable | 9.671 | 1 | **0.002 \*\*** | 8.874 | 1 | **0.003 \*\*** |
| Knowledgeable of any PCa risks factors | 19.032 | 1 | **0.001 \*\*** | 5.435 | 1 | **0.020 \*** |
| History of PCa in the family | 0.487 | 1 | 0.485 | 7.086 | 1 | **0.008 \*\*** |

(Asterisks indicate the level of significant differences: \*p ≤ 0.05; \*\*p ≤ 0.001; \*\*\*p ≤ 0.0001)

### Odds ratio on healthcare services and health seeking behavior towards prostate cancer screening

Odd ratio (OR) and 95% confidence interval (CI) were obtained for variables, which were found to be significant in “ever been screened” and “willingness to be screened” for PCa. Results from the odds ratio (OR) indicate that the respondents who were aware of PCa screening services were nineteen times more likely to ever been screened for prostate cancer (PCa) (OR=19.000; 95% CI, p=0.001). Similar to the awareness of whether PCa is manageable and knowledge of risk factors were more than thirteen times and six times likely to have been screened for prostate cancer (OR=13.571; 95% CI, p=0.001 and OR=6.714; 95% CI, p=0.001) respectively. Consulting a general medical practitioner was less likely (OR=0.087; 95% CI, p=0.99) associated with an individual’s intent to be screened for prostate cancer (PCa). However, awareness of PCa is manageable and knowledge of risk factors were more than five times and six times likely to influence individual participation in prostate cancer screening (OR=5.500; 95% CI, p=0.009 and OR=6.523; 95% CI, p=0.072) respectively. Additionally, the respondents who reported a history of PCa in the family were likely (OR=1.116; 95% CI, p=0.043) to uptake prostate cancer screening as shown in Table 5 below.

**Table 5.** Odds ratio of binary logistic regression on healthcare services and health seeking behavior among men aged 30-64 years old in Mukuru informal settlement.

| Variables | Ever been screened |  |  |  |
| --- | --- | --- | --- | --- |
|  | p Value | Odds ratio (OR) | 95% Confidence Interval |  |
|  |  |  | Lower | Upper |
| Aware of PCa screening services | **0.001\*\*** | 19.000 | 8.885 | 40.629 |
| Aware of any PCa screening method | 0.618 | 1.286 | 0.479 | 3.452 |
| Aware if PCa is manageable | **0.001\*\*** | 13.571 | 6.299 | 29.241 |
| Knowledge of any PCa risks factors | **0.001\*\*** | 6.714 | 3.035 | 14.854 |
| Variable | Willingness to be screened |  |  |  |
|  | p Value | Odds ratio (OR) | 95% Confidence Interval |  |
|  |  |  | Lower | Upper |
| Consulting general medical practitioner | 0.99 | 0.087 | 0.011 | 0.660 |
| Aware if PCa is manageable | **0.009\*\*** | 5.500 | 1.529 | 19.789 |
| Knowledge of any PCa risks factors | **0.072\*** | 6.523 | 0.844 | 50.437 |
| History of cancer in the family | **0.043\*** | 1.116 | 1.059 | 1.175 |

(Asterisks indicate the level of significant differences: \*p ≤ 0.05; \*\*p ≤ 0.001; \*\*\*p ≤ 0.0001)

**Table 6.** In-depth interview on utilization of PCa screening services among men aged 30-64 years old from Mukuru informal settlement in Kenya.

| Code | Categories | Strongly Emerging themes |
| --- | --- | --- |
| **Ever been tested** | Reasons for not been screened | • Never felt sick<br>• No idea where to get free PCa screening<br>• Not aware of PCa screening<br>• Lack of time and interest<br>• Ignorance<br>• Fear of unknown |
| **Willingness** | Reasons for willing to be screened of PCa in future. | • To know their health status<br>• To detect cancer at early stage<br>• Due to aging when they reach 40yrs & above<br>• Availability of free PCa screening in their nearest healthcare Centre. |
| **Not willing** | Reasons for not willing to be screened in future | • Fear of unknown<br>• Lack of time and interest<br>• Dislike of hospitals |

**Table 7.** Key informant Information on health care services and PCa screening uptake among men aged 30-64 years old from Mukuru informal settlement in Kenya.

| Code | Categories | Strongly emerging themes |
| --- | --- | --- |
| **Provision of PCa screening services** | Information concerning PCa screening | • Mostly done one-on-one during consultations<br>• Lack of PCa screening services<br>• Lack of awareness and the boy child empowerment |
|  | Availability of PCa screening equipment's and skilled workforce | • Available in very few private hospital at a cost<br>• Lack of On-job-training concerning PCa screening.<br>• Lack of PCa screening guidelines and standard operational procedures |
| **Policy** | How PCa screening policies can be improved to benefits low-income population | • Resource mobilization and allocation<br>• Partnering with donors / Institutions<br>• Community sensitization and awareness<br>• Proper strategic plan from MoH down to the healthcare facilities |
| **Implementation** | Effective ways to reach out, involve and motivates men to uptake PCa screening. | • Budget allocation for free PCa screening services<br>• Capacity building for health care workers and community health promoters<br>• Men sensitization on PCa & mobilization. |
| **Resources** | Availability of budgetary funds allocation or donor support specific to PCa screening | • Insufficient resources due to lack of funds.<br>• Lack of budget allocation to provide PCa services. |
| **Challenges** | Challenges facilities faced in provision of PCa screening. | • Lack of budget allocation to provide PCa services.<br>• Lack of proper implementation plan. |
|  | Challenges faced by men which lead to low utilization of PCa screening services. | • Cultural beliefs, Stigma, anxiety and fear of unknown.<br>• Poor health seeking behavior in men. |
|  | How to mitigate or reduce the challenges identified. | • Develop a program that capture men's attention and create interest for them to embrace prostate cancer screening. |

### Responses on utilization of health care services in regards to prostate cancer screening

During an in-depth interview with respondent’s majority strongly conveyed the following reasons to justify why they had not been screened for PCa; they had never felt sick to seek medical attention, they were unaware of male screening, lack of time and interest. They also expressed willingness to be screened in the future. A few (8.5%) respondents expressed their unwillingness to be screened due to fear of the unknown and lack of time among other reasons.

### In-depth interview with key informants on health care services provision and prostate cancer screening uptake

To obtain more information on factors influencing the uptake of prostate cancer screening among men in Mukuru informal settlements, the study conducted an in-depth interview with key informants who were mostly health care services providers and community health volunteers within the facilities in the study areas. The following information was obtained during in-depth interview with the key informants.

### Provision of PCa screening services

The majority of healthcare facilities in the study area do not provide PCa screening services due to a lack of testing kits and funds, only a few private facilities do offer prostate-specific antigen (PSA) testing at a cost. Concerning skills of healthcare workers most complained they lacked on-job training on PCa and screening guidelines. This was verified by an observational checklist where most of the facilities lacked screening equipment, PSA tests and there were no charts or fliers that displayed any information on PCa and screening. A couple of the service providers commented that “*the Ministry of Health and other health agencies have paid more attention to breast and cervical screening and forgotten about PCa screening”.* Information concerning PCa screening was not at the client’s disposal but was conveyed through one-on-one consultation only for suspicious cases where the patient was advised on screening for diagnostic purposes. Majority of clinicians referred these cases to either Mbagathi County Hospital or Kenyatta National Hospital.

### Prostate cancer screening policy

A couple of ideas emanated from the key informants from various health facilities on how PCa screening policies can be improved to benefit low-income populations, populations at risk and rural men. The majority of key informants were of the opinion that the government through the Ministry of Health (MoH) should partner with donors to fund the PCa screening programmes countywide, create awareness through community strategies and sensitization in a well-established strategic plan that will capture the low-income populations and rural men. Plan and coordinate a well-organized PCa screening programme from MoH down to the healthcare facilities and then to the communities targeting the low-income populations, populations at risk and rural men.

### Implementation of PCa screening services

The facilities in-charge and medical officers reiterated that the provision of free, accessible prostate cancer screening services and resource allocations were some of the most effective strategies to reach out, involve and motivate men to uptake PCa screening. In addition, the capacity building of healthcare workers and community health volunteers similar to other health programmes like in HIV/AIDS and T.B, cervical or breast cancer screening for women would improve the uptake and sensitization on PCa screening. The facilities in-charge, medical officers and clinicians indicated that there was a lack of a well-established allocation of funds from the Ministry of Health or donor support dedicated to the improvement of PCa screening in level 2&3 healthcare facilities which can be used to reach out to the low-income populations. Hence, there were no resources or budget allocations to public healthcare facilities within Mukuru informal settlements that had been set aside for the purpose of increasing the uptake of PCa screening.

### Challenges in providing PCa screening services

Almost all public healthcare facilities in Mukuru experienced challenges in provision of the services due to a lack of budget allocation to purchase PSA screening testing kits and equipment. The lack of a proper implementation plan was identified to be the main challenge facing most health facilities. Poor health-seeking behavior, cultural beliefs, stigma, anxiety and fear of the unknown were stated as the main challenges for the men. The majority of facilities in-charge and medical officers were of the opinion that the best way to address the challenges of low uptake of PCa screening is to partner with donors and other institutions/organizations in health sectors including the MoH in developing a strategic programme that will address issues of the men in seeking health care services.

## DISCUSSION

### Socio-demographic factors

The study found that age, marital status, occupation and household monthly income have no influence on the uptake of PCa screening. Similar to the findings were reported from a study conducted in western Kenya among the male healthcare workers found that age, marital status and religion not associated with PCa screening practices [20]. In addition, this was contrary to the findings reported in a study at Kitwe Teaching Hospital in Zambia where age above 60 years was associated with PCa screening practices and knowledge about the disease itself [21]. The differences in findings may be explained by that the participants in the present study who reported age between 60-64 years were very few (6.5%) compared to more than a quarter of participants who reported age above 60 years in the study at Kitwe Teaching Hospital in Zambia. However, this study found an association between the level of education and ever been screened for PCa among men in the Mukuru informal settlements. Low education levels significantly relate to unawareness of prostate cancer screening and high mortality rates globally. Knowledge comes along with education level and screening history is key to the willingness to undergo screening in the future [22]. Similar findings have been reported among men in Tanzania which showed that men with formal education were likely to access health information, internalize it and utilize health care services [23]. Similarly, a study on factors associated with PSA uptake among Australian men reported an increased odds ratio (OR) for men with higher education compared to those with no qualifications [24]. However, the odds of having a PSA test were higher in men who were married or living with a partner compared to unmarried or singles [24]. This was contrary to the findings from this study where marital status was not associated with PCa screening uptake among men aged 30-64 years in Mukuru informal settlements.

The monthly income of the respondents was not associated with PCa screening uptake among men in the Mukuru informal settlements. This can be explained by the fact that more than half of the respondents reported earning less than KES 15,000 per month. Thus most of them lacked medical insurance cover and were unable to pay for their medical expenses. During the in-depth interview, most of the respondents reported being unable to make enough for their basic needs. This is somehow similar to a study done in Nigeria which found that an increase in income increases the odds or likelihood of Nigerian men intent to be screened for PCa [25]. Due to the high level of risk involved and the requirement that employees respond to emergencies in challenging environments, careers such as firefighting and police work have been linked to an increased risk of cancer [26]. This affirms the need to understand risk factors in different occupation and their effect on prostate cancer screening. For instance, the findings from this study found occupation (source of income) was not significantly associated with an individual’s participation in PCa screening. This may be due to that most of the respondents were casual laborers and others self-employed in their small businesses hence they lacked the time and were not able to earn enough to pay for the PSA screening test. Therefore, if the local county government can implement a mobile clinic that offers free PSA screening to these men at their place of work may improve early detection and diagnosis, especially for the men who are at risk.

### Healthcare services and health seeking behavior on prostate cancer screening in Mukuru informal settlements

The results of this study show that most of the Mukuru residents were near to either public or private healthcare facilities reported a distance between 1 - 2 kilometers. Despite that, they had access to health care services these healthcare facilities lacked PSA screening tests and men’s sensitization on PCa screening. This differs in the rural areas where distance determines the utilization of a healthcare facility. In addition, the findings from a study done in Lira city- Uganda found that most of the participants reported the distance from their nearest health facility was between 5 kilometers [27]. Consulting of a medical practitioner was highly significantly associated with willingness to be screened for PCa in the future. For instance, most clinicians convey information about PCa through one-on-one consultation with the patients when it is necessary. However, due to the long queue, not all the male patients seen on that day were informed about PCa screening, as reported by one of the clinicians “*I only advise on PCa screening to male clients whose symptoms presenting similar to PCa for diagnosis purposes”.* This is an indication that informed decision-making about PCa screening was not provided or available to all the male clients visiting the healthcare facilities hence men were less likely to uptake PCa screening which resulted to low uptake within Mukuru informal settlements. A similar result was reported from a study conducted in a rural community in Kiambu-Kenya where 10% of screened men had a recommendation from a healthcare provider but most of them were not involved in informed decision-making [6]. This shows lack off or very little is done on men sensitization on PCa and screening within healthcare facilities. In addition, a study done on PCa screening communication among young men of different ethnic groups reported a gap on racial disparities in shared decision-making between non-Hispanic (black men) and medical practitioners thus they were less likely to be involved in informed decision-making regarding PCa and PSA screening as compared to Hispanic white men [28]. This is in contrast to studies done in developed countries where family medical doctors were the main source of information about PCa and screening resulted in a high level of PCa screening uptake. Therefore, there is a need for capacity building for our healthcare workers, the printing of posters, pamphlet, magazines, and even the use of local media services to convey information about PCa and screening domains.

The study also found that most of the respondents had no insurance cover and they paid their medical expenses from their own pocket. This affected individual decision-making on whether to pay for the PSA test or buy food for their family since most of them earned from their daily hustle. For instance, during an in-depth interview, one of the respondents reported that the reason why he had never been screened was that; “*the service was not provided in the healthcare center so I went to a private facility where I was charged for 1000ksh and I didn’t have sufficient funds so I never took the test”*. This is contrary to some studies done among Nigerian men where most of the participants were comfortable paying for their medical expenses where the majority of them had medical insurance and others had high-paying jobs with adequate cover [29,30]. It is likely that payment did not reflect as a barrier in the present study because most respondents were not aware of PCa screening services or the actual costs of the service and most of the healthcare facilities in Mukuru informal settlements lacked PSA testing. Based on the report [13], primary healthcare services are not equally accessible, affordable and available in countries in Southern Asia and Sub-Saharan Africa. The burden of prostate cancer in developing nations is mostly a result of limited access to services such as screening, prevention and control [13]. Therefore, this calls for Local governments and MoH to consider providing free PCa screening services and equipment accompanied by capacity building for healthcare providers to all healthcare facilities that serve low-income populations and in rural areas inorder to increase utilization of PCa screening. For instance, during an in-depth interviews with health care providers reported that: “*they lacked PSA testing kit for screening men, mostly what is offered are breast and cervical screening, TB and HIV/AIDS testing that has been budgeted for.”* Moreover, on the issues of policy and implementation of PCa screening services, the facility in-charges and medical officers reiterated that the provision of free and accessible screening services together with resource allocations were some of the most effective strategies to reach out, involve and motivate men to take up PCa screening. One of the key informants reported that “*the main challenges that service providers face is lack of budgetary allocation for PCa screening, sensitization and poor health-seeking behavior in men who rarely go to hospitals”.* This indicates the need for MoH to collaborate with other parties in the health sector to develop a programme that can capture men’s attention and create interest for them to embrace prostate cancer screening.

The findings on health-seeking behavior of men residing in Mukuru informal settlements, the study found that most of the participants had heard about prostate cancer but were not aware of men’s screening services, and screening methods. Also they showed low knowledge level on predisposing risk factors for prostate cancer (PCa). Similar findings were reported from the work of [31] who found that the level of awareness of Kenyan men on PCa was good (61.9%) but low (3.9%) on PCa screening practices. Kenya continues to have extremely poor screening uptake, much like other underdeveloped nations. The level of participation is still low nationally despite the fact that Kenya’s current guidelines advocate screening men aged 40 to 69 years through an informed shared decision-making process [5]. The differences may be explained by a gap in information about PCa screening, its benefits, harm and risks. This calls for the MoH to improve the approaches in health communication and behavior change communication targeting men aged 40 and above. However, similar findings on information disparities were reported in several studies that assessed knowledge, awareness and practices towards PCa screening in Sub-Sahara African countries [32; 21 & 27]. Low knowledge level on PCa screening domains may have contributed to the low uptake of PCa screening where only a few (3.5%) of the respondents reported having been screened for prostate cancer by the time of the study. A similar finding was reported in a study conducted among men in Dzingahe village, Limpopo province where respondents had a positive attitude and were willing to uptake PCa screening but only a few (3.3%) had undergone screening [33]. Additionally, a study conducted among commercial motorcyclists in Nigeria found that less than one-fifth of the men had undergone screening [34].

Such evidence indicates there is still a persistent low PCa screening uptake in Sub-Sahara African countries suggesting poor health-seeking behavior among African men. During an in-depth interviews majority of the respondents conveyed that; “*the reasons why they had not been screened for PCa was that they had never felt sick to seek medical attention and they were unaware (critical awareness not just health seeking behavior) of men screening*”. Others reported that lack of time and interest were the main reasons why they had not gone for PCa screening. This indicates that it is important these services be brought closer and made conveniently as possible. An example of this is how the MoH provided HIV testing conveniently through night mobile clinics to key populations such as prostitutes, gays and injecting drug users. For instance, most of the participants were willing to uptake PCa screening in the future for the following reasons: “*to be aware of their health well-being, availability of free PCa screening services and detection of PCa status at an early stage which make it manageable”*. The study also found that a few who were not willing to uptake PCa screening was due to fear of the unknown, avoiding conventional medicine, lack of time and interest. Similar findings were reported in a study done in Kasekeu a rural area in Makueni County- Kenya, which found that African men held a high fatalistic beliefs, fear and apprehension concerning PCa and screening compared to African Americans who had a small degree of fear and fatalistic beliefs [35]. The difference could be due to the African American man being more informed about PCa and screening compared to his counterpart based in Africa. The MoH and Local Governments must consider extending accessible free prostate cancer screening programmes in healthcare facilities as well as enhance the application of modernized screening procedures that would encourage men to go for early PCa screening and diagnosis.

Knowledge of the risk factors and awareness that PCa is manageable were highly significantly associated with “ever been screened” and “willingness to be screened”. Awareness of screening methods showed a significant association with ever being screened for prostate cancer, this indicates that knowledge and awareness are associated with an increase in the uptake of prostate cancer screening. This underscores the importance of awareness of health information about PCa screening in influencing individual’s participation in the future. There is therefore the need to develop programmes that increase men’s knowledge on PCa, the importance of early detection and prostate cancer as a disease that is treatable at early stage, this may improve men’s health-seeking behavior. Therefore, this calls for vibrant health education and exposure to increase critical awareness and knowledge about PCa and screening. Additionally, history of PCa in the family was found to be highly associated with an individual’s willingness to screen for PCa. This might be most likely due to their past experiences in witnessing someone suffering from cancer.

Similarly, the work of [36] found that men who have a family history of PCa with inheritance gene mutation are expected to be more susceptible to the disease and are more likely to undergo screening. The study also found that genetic status influences the participants’ perception of prostate cancer risk [36]. In addition, a similar finding was reported in Rwanda where some of the participants who had a family history of PCa reported this as the reason why they consulted urologist services [37]. This indicates that men with a history of PCa in the family were more likely to seek PCa screening compared to men with no family history of PCa.

## CONCLUSION

The following conclusions are drown from the current study:

The socio-demographic factors like age, marital status, household monthly income and source of income (occupation) were not associated with the uptake of PCa screening among men of low-income settings. However, respondents who had formal education were more likely to have higher individual understanding, acceptance and utilization of the screening programme. This means that people may not prioritize the services because of cost and the fact that they feel healthy.

Primary health care providers play a key role in the uptake of PCa, as they are heavily involved in the health care system, that most of the variables assessed under healthcare services showed significant association in influencing individual participation in PCa screening uptake. Therefore, there is a need for health workers to be well-facilitated and equipped in order to help in addressing low PCa screening uptake in the country. Moreover, the implementation of modernized and friendly screening services may prevent men from shying away from screening and motivate them to uptake PCa screening. In addition, on the health-seeking behavior awareness of the predisposing risk factors, awareness of screening domains and aware of PCa is manageable at an early stage all are important determinants for the individual’s intent to uptake of PCa screening in the future.

## RECOMMENDATIONS

From the findings of this research, the following recommendations are suggested;

i. The local county government to partner with the MoH in creating critical awareness on the importance of routine PCa screening practices by ensuring that men are well-informed on screening domains, benefits, harms and risks.
ii. Provision of free modern PCa screening services and increasing accessibility to all public and private healthcare facilities that offer primary healthcare services especially in low-resourced areas.
iii. The MoH to establish a health programme that will regularly monitor men’s practices towards PCa screening uptake from the National level down to the Counties and sub-counties levels.

## Data Availability

All data produced in the present study are available upon reasonable request to the authors

## Consent for Publication

Not applicable

## Authors contributions

K.A as the principal author conceived, designed the study and acquired the data. Drs. E.M, F.O and J.N assisted in developing the methods, reviewing, analyzing and interpreting the data. All authors revised the manuscript.

## Principle Author’s information

K.A post-graduate student in Technical University of Kenya (TUK) in the department of health system management and public health.

## Acknowledgements

First, I take this opportunity to give thanks to the Almighty God for allowing me to go this far. I am indebted to my colleagues Mr. James Oula and Nancy Mitalo for their great insights, encouragement and intellectual guidance. Furthermore, I would like to acknowledge the healthcare workers and community units in Embakasi south constituency and Makandra constituency for their contributions to this study. For their steadfast support and efforts, I would like to thank the entire Technical University of Kenya (TUK) post-graduates research forum. I also like to thank the department’s staff and professors for their assistance in many aspects of various ways. Last but not least, I want to convey my sincere appreciation to my entire family for their ongoing support on all front.

